# Longitudinal lung function assessment of patients hospitalised with COVID-19 using ^1^H and ^129^Xe lung MRI

**DOI:** 10.1101/2022.04.06.22272747

**Authors:** Laura C Saunders, Guilhem J Collier, Ho-Fung Chan, Paul J C Hughes, Laurie J Smith, James Watson, James Meiring, Zoë Gabriel, Thomas Newman, Megan Plowright, Phillip Wade, James A Eaden, Jody Bray, Helen Marshall, David J Capener, Leanne Armstrong, Jennifer Rodgers, Martin Brook, Alberto M Biancardi, Madhwesha R Rao, Graham Norquay, Oliver Rodgers, Ryan Munro, James E Ball, Neil J Stewart, Allan Lawrie, Gisli Jenkins, James Grist, Fergus Gleeson, Rolf F. Schulte, Kevin M Johnson, Frederick Wilson, Anthony Cahn, Andrew J Swift, Smitha Rajaram, Gary H Mills, Lisa Watson, Paul J Collini, Rod Lawson, A A Roger Thompson, Jim M Wild

## Abstract

**Introduction:** Microvascular abnormalities and impaired gas transfer have been observed in patients with COVID-19. The progression of pathophysiological pulmonary changes during the post-acute period in these patients remains unclear.

**Methods:** Patients who were hospitalised due to COVID-19 pneumonia underwent a pulmonary ^1^H and ^129^Xe MRI protocol at 6, 12, 25 and 51 weeks after hospital admission. The imaging protocol included: ultra-short echo time, dynamic contrast enhanced lung perfusion, ^129^Xe lung ventilation, ^129^Xe diffusion weighted and ^129^Xe 3D spectroscopic imaging of gas exchange.

**Results:** 9 patients were recruited and underwent MRI at 6 (n=9), 12 (n=9), 25 (n=6) and 51 (n=8) weeks after hospital admission. Patients with signs of interstitial lung damage at 3 months were excluded from this study. At 6 weeks after hospital admission, patients demonstrated impaired ^129^Xe gas transfer (RBC:M) but normal lung microstructure (ADC, Lm_D_). Minor ventilation abnormalities present in four patients were largely resolved in the 6–25 week period. At 12 week follow up, all patients with lung perfusion data available (n=6) showed an increase in both pulmonary blood volume and flow when compared to 6 weeks, though this was not statistically significant. At 12 week follow up, significant improvements in ^129^Xe gas transfer were observed compared to 6-week examinations, however ^129^Xe gas transfer remained abnormally low at weeks 12, 25 and 51. Changes in ^129^Xe gas transfer correlated significantly with changes in pulmonary blood volume and TL_CO_ Z-score.

**Conclusions:** This study demonstrates that multinuclear MRI is sensitive to functional pulmonary changes in the follow up of patients who were hospitalised with COVID-19. Impairment of xenon transfer may indicate damage to the pulmonary microcirculation.

## Introduction

In patients hospitalised with pneumonia due to SARS-CoV-2 infection, the existing literature and clinical experience suggest that there is considerable overlap in clinical presentation with typical pneumonia and acute respiratory distress syndrome (ARDS), with patients exhibiting hyper-inflammation and progressive hypoxaemia. However, patients with severe COVID-19 also show evidence of an inflammatory and thrombotic vasculopathy with endothelial dysfunction and excessive blood flow to collapsed lung tissue[1-3]. Abnormal pulmonary vasoregulation has been observed in patients in the acute phase of COVID-19[1] and may be a pathophysiological mechanism contributing to the progressive hypoxemia seen in these patients. The medium and long-term outcomes and complications of microvascular abnormalities alongside pulmonary scarring in patients who have been hospitalised due to COVID-19 are currently unknown.

Lung MRI with hyperpolarised ^129^Xe gas allows direct, regionally sensitive measurements of lung ventilation and gas diffusion within the lung airspace (diffusion weighted MRI, DW-MRI). DW-MRI provides a quantitative assessment of acinar airway dimensions. The derived apparent diffusion coefficient (ADC) and mean acinar airway dimensions (LmD) provides 3D in vivo information of the underlying lung microstructure, and has been shown to be highly sensitive to changes in lung microstructure in patients with emphysema[4] and fibrotic lung disease[5].

In addition, ^129^Xe is soluble in the lung tissue membrane (M) and in the red blood cells (RBC), and the signal from ^129^Xe in these dissolved compartments can be distinguished spectroscopically. The ratio of the ^129^Xe MRI signal observed in the lung airspaces (gas), the lung membrane and bound to the red blood cells can thus be determined with magnetic resonance spectroscopic imaging. In particular, the fraction of the ^129^Xe signal in the RBC:M, RBC:gas and M:gas have been used to probe the transfer of gas between the airspace, membrane and blood[6, 7]. RBC:M has been shown to be highly sensitive to gas transfer limitation and longitudinal assessment of change in interstitial, emphysematous and pulmonary vascular diseases[8-10].

An early study from Wuhan, China with ^129^Xe MRI showed reduced gas transfer to the RBC in 13 patients scanned 14-34 days after discharge from hospital following acute COVID-19, and a small but significant increase in lung ventilation defects when compared to healthy subjects[11]. Reduced xenon gas transfer has been shown to persist in patients hospitalised with COVID-19 pneumonia 24 weeks after discharge from hospital and it has been suggested that this may provide an explanation for prolonged breathlessness (a Long-COVID symptom) in patients[12]. In 11 non-hospitalised Long-COVID patients and 12 post-hospitalised long-COVID patients, RBC:M was reported to be significantly decreased compared to healthy volunteers[13] and low RBC:M has been reported in hospitalised patients 34 weeks after infection compared to controls[14]. However none of these studies had controls which were age and sex-matched to the patient cohorts, and it has been suggested that age related differences are likely to be contributing to the decreased RBC:M ratio reported in some studies into COVID-19 patients[15]. It is currently unclear what is driving the decreased xenon gas-transfer reported in patients who have had COVID-19. As xenon gas transfer depends on both the xenon uptake in the lung tissue and the xenon uptake in the red blood cells, lung perfusion abnormalities or alveolar/interstitial endothelial changes, or a combination of both, may be driving the reduced xenon gas transfer seen in patients after COVID-19. Lung perfusion imaging has been proposed as a key triage tool to evaluate small vessel injury and residual blood clot in patients who have had COVID-19[16]. Dynamic contrast enhanced (DCE) ^1^H lung MRI allows the assessment of lung perfusion without exposing the participant to ionising radiation and is therefore well suited to patient follow-up studies. In one patient who was hospitalised due to COVID-19, 44% of the lung volume demonstrated a delayed or absent contrast signal-enhancement peak[17]. In another study of 28 people with persistent dyspnea after COVID-19, patients demonstrated significantly longer mean pulmonary time-to-peak (TTP) when compared with healthy controls (preprint)[18]. This evidence suggests lung perfusion MRI has sensitivity to detect microvascular perfusion abnormalities in patients who have had COVID-19.

Furthermore, ^1^H lung MRI can also provide structural lung images with good visualisation of the lung parenchyma using ultra-short echo time (UTE) imaging. UTE imaging has been reported to be of comparable quality to CT imaging for evaluating lung structural changes due to COVID-19 in a cohort of 23 acute COVID-19 patients[19].

In this study we utilised a comprehensive, multinuclear MRI protocol which combines hyperpolarised ^129^Xe imaging methods sensitive to ventilation, lung microstructure (DW-MRI) and gas exchange (dissolved xenon spectroscopic imaging) alongside ^1^H DCE perfusion and UTE lung structural imaging to assess pathophysiological changes in patients who had been hospitalised with COVID-19 pneumonia, during the post-acute period.

## Methods

### Participants

Patients with acute COVID-19 pneumonia and no previously diagnosed respiratory disease (excluding mild asthma) were recruited from Sheffield Teaching Hospitals respiratory and infectious diseases wards from November 2020. Recruitment for this study is ongoing, with follow-up ^29^Xe and ^1^H lung MRI examinations at approximately 6, 12, 24 and 52 weeks after COVID-19 infection.

Patients were required to meet the following criteria:

i. a positive SARS-CoV-2 result from a nasal/pharyngeal or respiratory sample;
ii. hospitalisation with a diagnosis of pneumonia (chest X-ray or CT scan consistent with COVID-19 infection);
iii. development of impaired oxygenation (SpO2≤93% on room air) requiring additional oxygen
iv. did not show evidence of interstitial lung damage on CT or MRI structural imaging at 12 weeks after hospital admission, as judged by a clinical chest radiologist.

Patients with evidence of interstitial lung damage at 12 weeks after hospital admission were recruited into the parallel UKILD study[20]. Standard MRI exclusion criteria were applied to all subjects. In addition, patients were excluded if they were unable to tolerate a test inhalation of ^129^Xe gas according to supervising clinicians’ judgement, or if they had a chest size exceeding the ^129^Xe chest coil diameter (38 cm). A flow diagram of patient recruitment is shown in Figure 1.

**Figure 1:**
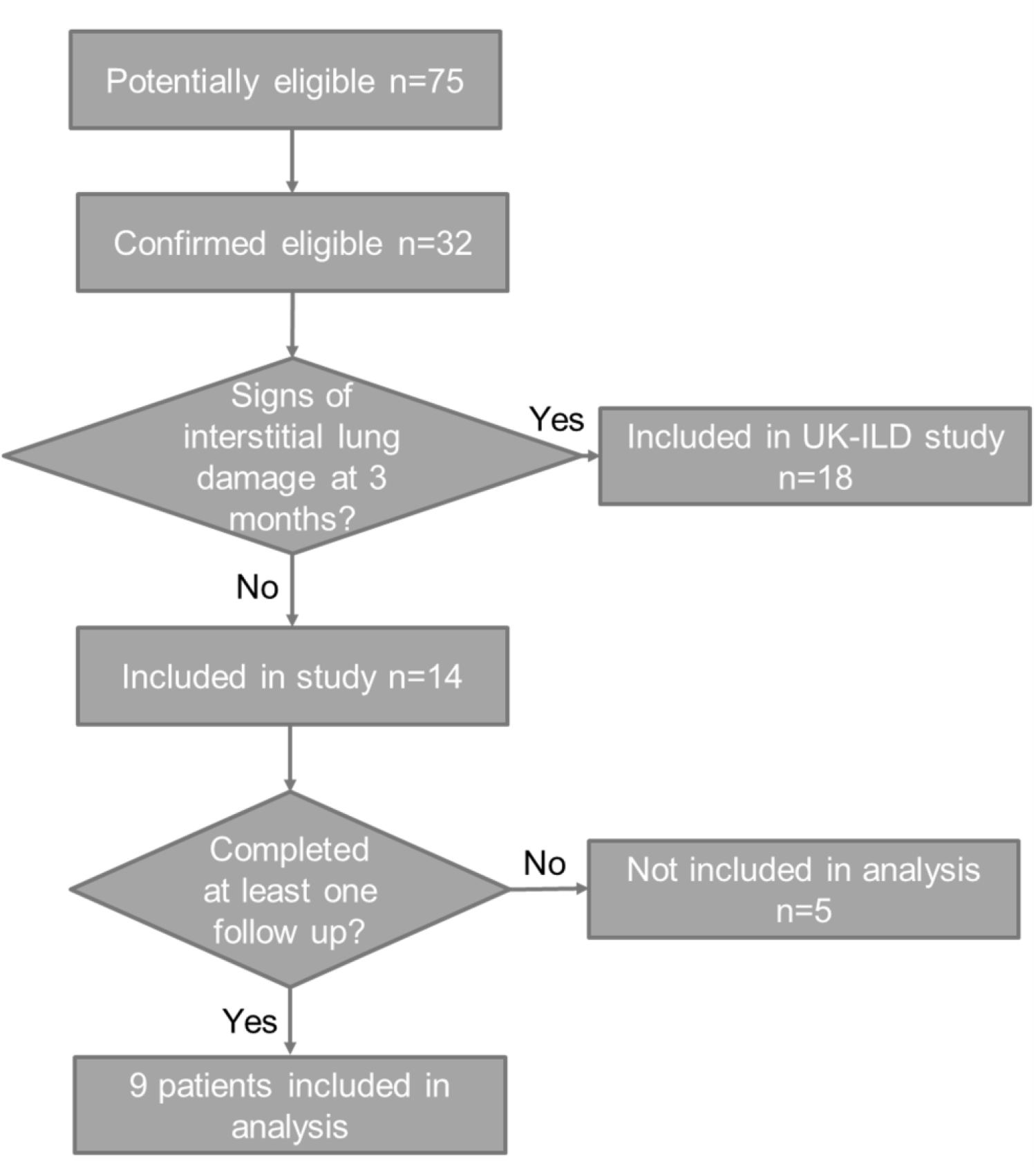
A flow diagram detailing patient recruitment into this ongoing study. Patients with signs of interstitial lung damage at 12 weeks were recruited into UKILD[20].

### MRI acquisition

Patients underwent scanning on either a GE HDx 1.5T (N=7) or a GE 450W 1.5T (GE Healthcare, Milwaukee, WI, USA) (N=2) MRI scanner[21], that is regulatory approved for manufacture of hyperpolarized ^129^Xe for clinical lung MRI by the UK MHRA. Each patient underwent MRI examinations on the same scanner for baseline visits and follow-up. See Figure 2 for an illustrative diagram of lung MRI methods utilised within this study.

**Figure 2:**
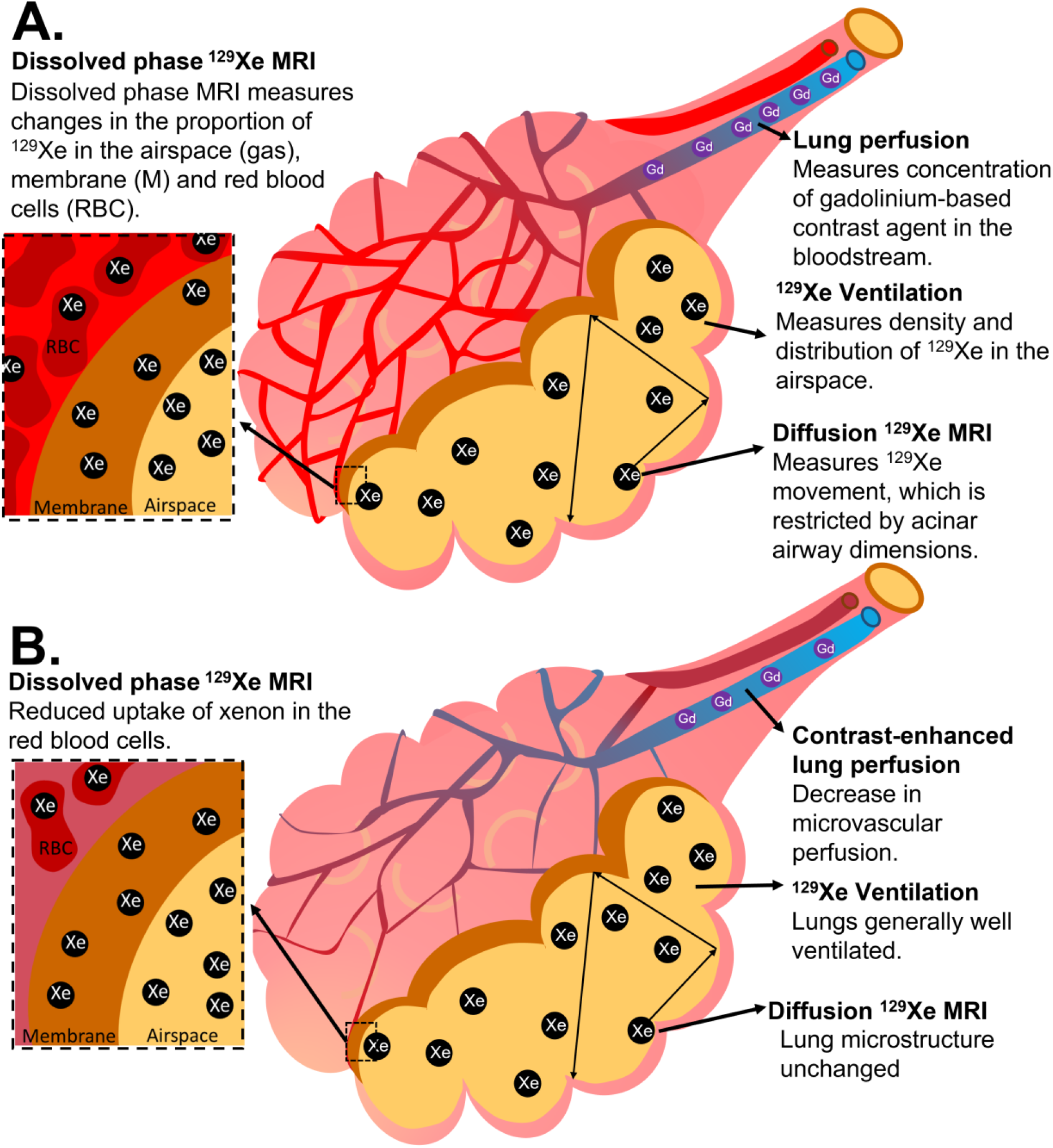
Illustrative diagram showing how the lung MRI techniques employed in this paper measure to lung perfusion, ventilation, lung microstructure (acinar airway dimensions) and xenon gas transfer (the transfer of xenon between the airspace, membrane, and RBCs). A) shows these techniques in a healthy alveolus. B) shows a possible interpretation of the findings of this paper in patients who have had COVID-19, with reduced RBC:M due to damage to pulmonary microcirculation but preserved acinar airway dimensions.

^129^Xe doses were polarised to ∼30% using a home-built high performance spin-exchange optical pumping polariser[21]. ^129^Xe images were acquired with the patient in a flexible quadrature transmit/receive vest coil (Clinical MR Solutions, Brookfield, Wisconsin, USA). Patients’ vital signs were monitored throughout the MRI examination. MR imaging parameters are detailed in the online supplement, Supplementary Table 1.

A structural ^1^H scan was acquired after inhalation of a 1L bag of air to match the lung inflation state of the subsequent xenon sequences. ^129^Xe ventilation images were acquired after inhalation of a 1L mixture of ^129^Xe and N_2_ using a 3D imaging sequence[22] detailed in the online supplement, Supplementary Table 1.

^129^Xe DW-MRI to assess alveolar micro-structural change was acquired after inhalation of 1L mixture of ^129^Xe and N_2_ using a 3D spoiled gradient echo (SPGR) multiple b-value sequence with compressed sensing[23].

3D spectroscopic imaging of the gas and dissolved phase xenon resonances (dissolved xenon in lung membrane, M, and in blood red blood cells, RBC) was acquired using 1L of hyperpolarised ^129^Xe[24].

^1^H MRI was acquired using an 8-element cardiac array (GE Healthcare, Milwaukee, WI, USA). UTE images were acquired with a 3D radial sequence during 8 minutes of free-breathing with prospective respiratory bellows gating on expiration[25].

3D variable flip angle SPGR images[26, 27] were acquired with flip angles of 2°, 4°, 10° and 30°, to allow for the correction of lung T_1_ and proton density. DCE lung perfusion MRI was acquired using time-resolved SPGR imaging with view sharing and parallel imaging. A half dose (0.05mL/kg) of Gadovist (Bayer) was administered at an injection rate of 4ml/s followed by a 10ml saline flush at 4ml/s. Patients were advised to hold their breath for as long as possible and breathe shallowly thereafter.

### Image analysis

Qualitative assessments of the UTE ^1^H structural, ^129^Xe ventilation and DCE lung perfusion images were made by two radiologists with 10 and 14 years’ experience. UTE images were assessed for parenchymal changes, and ventilation and perfusion images were assessed for defects.

Metrics of ventilation defect (VDP), low ventilation percentage (VP), normal VP, and hyper VP for each patient were calculated using linear binning (see online supplement). The coefficient of variation (CV) of the segmented lung ventilation images was also calculated from the ^129^Xe ventilation images as metrics of ventilation heterogeneity.

Maps of ^129^Xe apparent diffusion coefficient (ADC) and mean diffusive length scale (Lm_D_) from a stretched exponential model of ^129^Xe gas diffusion in the lungs were calculated on a voxel-by-voxel basis[28].

Maps of gas transfer ratios (RBC:M, RBC:gas, M:gas) were calculated from 3D spectroscopic imaging. The transverse relaxation time, T_2_^*^, of the RBC and M spectroscopic peaks was also calculated.

Mean values of all global metrics were calculated for each patient.

### Pulmonary function tests

Where possible, pulmonary function tests (PFTs) were acquired on the same day as the MRI examination at each visit. Spirometry and transfer factor were performed and from these tests, the metrics FEV1, FVC, FEV1/FVC, TL_CO_ and K_CO_ were calculated and presented as Z-scores and % predicted using GLI reference ranges[29, 30].

### Statistical analysis

Global MRI metrics from visits 1, 2, 3 and 4 were compared using a Skillings-Mack test due to the presence of missing data[31] with pairwise Wilcoxon tests and a correction for multiple testing was applied[32] using R software[33]. Data are presented as median (range), unless otherwise stated.

The within-subject correlation between RBC:M and i) DCE metrics and ii) TL_CO_Z was calculated using the analysis of covariance method[34] using IBM SPSS Statistics 27 (SPSS, New York, USA).

A p-value of <0.05 was considered statistically significant.

### Age and sex matched healthy volunteer metrics

Median ADC and Lm_D_ values for an age and sex matched control cohort were determined by retrospective analysis of previously published data[35]. 11 subjects from this previously published work were selected based on age and sex from a cohort of 23 whilst blinded to MRI metrics, such that the control cohort had a median age of 63 (40-70) years and 27% were female.

Median RBC:M, RBC:gas and M:gas for an age and sex matched control cohort were determined by retrospective analysis of a healthy cohort data set, with controls chosen based on age and sex whilst blinded to MRI metrics. 12 subjects were selected who had a median age of 57 (41-68) years and 33% were female.

## Results

Of the 75 patients approached, 32 were recruited. 5/75 approached participants were excluded due to their chest size exceeding the size of the xenon MRI coil and 2/75 approached participants were excluded due to poor health. Figure 1 shows a flow diagram of the number of individuals at each stage of the study, and further details on reasons for non-participation in the study are available in the online supplement. Of the 32 recruited patients, 14 did not show any signs of interstitial lung damage at 12 weeks and were therefore included as part of this study. 9/14 patients had follow-up examinations and were therefore included in the analysis in this paper.

3/9 patients were female. Median (range) patient age, height and weight were 57 (42-72) years, 173 (170-191)cm and 101 (84-112)kg. Visit 1 (n=9) occurred 6 (4-12) weeks after hospital admission, visit 2 (n=9) occurred 12 (11-22) weeks after hospital admission, visit 3 (n=7) occurred 25 (23-28) weeks after hospital admission and visit 4 (n=8) occurred 51 (49-62) weeks after hospital admission. Patients had been admitted to hospital with COVID-19 for 6 (2-15) days. Further patient demographic data are available in Table 1.

**Table 1.** Patient demographic data. MRC: medical research council. Ex-smokers all had ≤15 pack years.

|  | Group | Number of patients |
| --- | --- | --- |
| Age | <50 | 2 |
|  | 50-59 | 3 |
|  | 60-69 | 3 |
|  | 70-79 | 1 |
| Sex | Male | 6 |
|  | Female | 3 |
| BMI | 25-29.9 | 3 |
|  | 30-39.9 | 5 |
|  | 40+ | 1 |
| Oxygen requirement during hospital admission | Low flow | 8 |
|  | CPAP | 1 |
| ISARIC 4C score[37] | 1-4 | 3 |
|  | 5-8 | 5 |
|  | >8 | 1 |
| Length of stay | 1-5 | 4 |
|  | 6-9 | 4 |
|  | >10 | 1 |
| MRC Dyspnoea Scale (1-5) – 6 weeks | 1 | 6 |
|  | 2 | 2 |
|  | Not available | 1 |
| MRC Dyspnoea Scale (1-5) – 3 months | 1 | 7 |
|  | 2 | 1 |
|  | 3 | 1 |
|  | Not available | 0 |
| Comorbidities 4C score[37] | 0 | 4 |
|  | 1 | 4 |
|  | 3 | 1 |
| Smoking History | Never smokers | 4 |
|  | Ex-smokers | 5 |

UTE and ^129^Xe MRI were successfully acquired in all patients at all visits. DCE lung perfusion imaging was successfully acquired in 6/9 patients at visit 1, 8/9 patients at visit 2, 6/7 patients at visit 3 and 5/8 patients at visit 4, due to patient difficulties with the required breath hold for this acquisition.

Figure 3 shows representative slices from the UTE images, RBC:M maps, ^129^Xe ventilation images, and DCE pulmonary blood flow maps for each patient at visit 1 and visit 2. Figure 4 shows plots of ventilation, dissolved phase ^129^Xe, and DCE lung perfusion metrics for each patient at each visit. Median metrics and statistical comparisons of metrics at each visit are shown in Table 2.

**Table 2.** Median metrics for all MRI parameters at visits 1, 2, 3 and 4. Data are presented as median (range) of all patients with available data for each visit. If a Skillings-Mack test determined that there was a significant difference between at least two variables, p-values are shown for Wilcoxon pairwise tests. P-values are shown before and after adjustment for multiple testing.

|  | Visit 1 | Visit 2 | Visit 3 | Visit 4 | P-values | Adj P-values |
| --- | --- | --- | --- | --- | --- | --- |
| **N** | **9** | **9** | **7** | **8** |  |  |
| **ADC (cm<sup>2</sup>s<sup>-1</sup>)** | 0.0344<br>(0.309-0.0373) | 0.0327<br>(0.0281-0.0386) | 0.0340<br>(0.310-0.364) | 0.0338<br>(0.307-0.0357) | - |  |
| **Lm<sub>D</sub> (μm)** | 281 (260-300) | 273 (251-301) | 278 (263-290) | 279 (263-288) | - |  |
| **RBC:M** | 0.22 (0.15-0.37) | 0.25 (0.18-0.41) | 0.25 (0.19-0.44) | 0.23 (0.19-0.44) | V1-V2,<br>p=0.004<br>V1-V3,<br>p=0.047<br>V1-V4,<br>p=0.039 | V1-V2,<br>p=0.023*<br>V1-V3,<br>p=0.094<br>V1,V4,<br>p=0.094 |
| **RBC:gas** | 0.0026<br>(0.0018-0.0039) | 0.0030<br>(0.0019-0.0038) | 0.0026<br>(0.0020-0.0040) | 0.0024<br>(0.0017-0.0038) | - |  |
| **M:gas** | 0.0113<br>(0.0091-0.0125) | 0.0114<br>(0.0081-0.0179) | 0.0101<br>(0.0091 – 0.0113) | 0.0094<br>(0.0082-0.0104) | V1-V4, p=0.031<br>V2-V4,<br>p=0.023<br>V3-V4,<br>p=0.031 | V1-V4,<br>p=0.063<br>V2-V4,<br>p=0.063<br>V3-V4,<br>p=0.063 |
| **M T<sub>2</sub><sup>\*</sup> (ms)** | 2.58 (2.46-2.68) | 2.47 (2.38-2.58) | 2.42 (2.36-2.56) | 2.22 (1.94-2.40) | V1-V2,<br>p=0.044<br>V1-V3,<br>p=0.023<br>V1-V4,<br>p=0.008<br>V2-V4,<br>p=0.008<br>V3-V4,<br>p=0.031 | V1-V2,<br>p=0.053<br>V1-V3,<br>p=0.031*<br>V1-V4,<br>p=0.023*<br>V2-V4,<br>p=0.023*<br>V3-V4,<br>p=0.047* |
| **RBC T<sub>2</sub><sup>\*</sup> (ms)** | 2.20 (2.05-2.48) | 2.16 (2.01-2.49) | 2.16 (2.06-2.32) | 2.27 (2.10-2.47) | - |  |
| **DCE PBV (ml/100ml)** | 37.8 (11.7-53.5) | 47.6 (15.0-60.2) | 45.3 (36.3-58.4) | 53.8 (52.46-60.72) | - |  |
| **SD PBV (ml/100ml)** | 18.0 (7.5-28.5) | 21.3 (13.0-24.8) | 23.8 (17.9-25.7) | 21.1 (20.8-22.0) | - |  |
| **IQR PBV (ml/100ml)** | 25.1 (7.8-34.9) | 27.8 (10.5-36.1) | 35.9 (24.2-41.5) | 29.5 (27.0-31.9) | - |  |
| **DCE PBF (ml/100ml/min)** | 76.9 (19.6-107.2) | 90.2 (30.7 – 109.5) | 71.1 (60.3-117.7) | 98.3 (93.4-116.2) | - |  |
| **SD PBF (ml/100ml/min)** | 45.8 (11.5-58.8) | 54.5 (32.0-75.0) | 48.6 (35.6-69.5) | 54.2 (41.0 – 65.4) | - |  |
| **IQR PBF (ml/100ml/min)** | 54.0 (14.1-61.3) | 59.2 (25.5-102.9) | 59.0 (43.2-78.7) | 64.8 (54.2 – 72.4) | - |  |
| **Median MTT (s)** | 6.5 (5.6-8.0) | 7.3 (6.4-8.0) | 7.1 (3.5-9.9) | 6.9 (2.3 – 7.6) | - |  |
| **SD MTT (s)** | 1.3 (0.9-1.6) | 1.2 (0.6-2.6) | 1.3 (0.5-2.2) | 0.7 (0.6-0.9) | - |  |
| **IQR MTT (s)** | 1.3 (1.1-2.0) | 1.6 (0.7-3.1) | 1.3 (0.6-3.5) | 0.7 (0.5-1.1) | - |  |
| **VDP (%)** | 1.6 (0.6-3.9) | 1.3 (0.7-2.6) | 1.2 (0.4-2.1) | 0.8 (0.4-3.7) | V1-V3,<br>p=0.016 | V1-V3,<br>p=0.094 |
| **Normal VP (%)** | 76.4 (62.5-77.7) | 76.9 (72.3-86.2) | 78.9 (67.4-81.5) | 81.1 (69.0 – 82.6) | V1-V2,<br>p=0.027<br>V1-V3,<br>p=0.031 | V1-V2,<br>p=0.093<br>V1-V3,<br>p=0.093 |
| **Low VP (%)** | 12.5 (10.0-15.4) | 11.9 (8.9-13.1) | 10.9 (9.1-14.8) | 10.6 (10.0-13.7) | - |  |
| **Hyper VP (%)** | 11.7 (9.5-18.3) | 11.0 (4.2-13.3) | 9.7 (7.9-15.8) | 8.4 (6.8 – 15.2) | - |  |
| **Lung ventilation CV (%)** | 29.0 (27.5-37.1) | 28.8 (22.3-32.2) | 26.9 (25.6-34.1) | 26.1 (25.0 - 33.3) | V1-V2,<br>p=0.040<br>V1-V3,<br>p=0.016<br>V1-V4,<br>p=0.040 | V1-V2,<br>p=0.078<br>V1-V3,<br>p=0.078<br>V1-V4,<br>p=0.078 |

ADC: apparent diffusion coefficient. Lm<sub>D</sub>: mean diffusive length scale. RBC: red blood cell. M: membrane. DCE: dynamic contrast enhanced. PBV: pulmonary blood volume. PBF: pulmonary blood flow. MTT: mean transit time. VDP: ventilation defect percentage. VP: ventilation percentage. CV: coefficient of variation.

**Figure 3:**
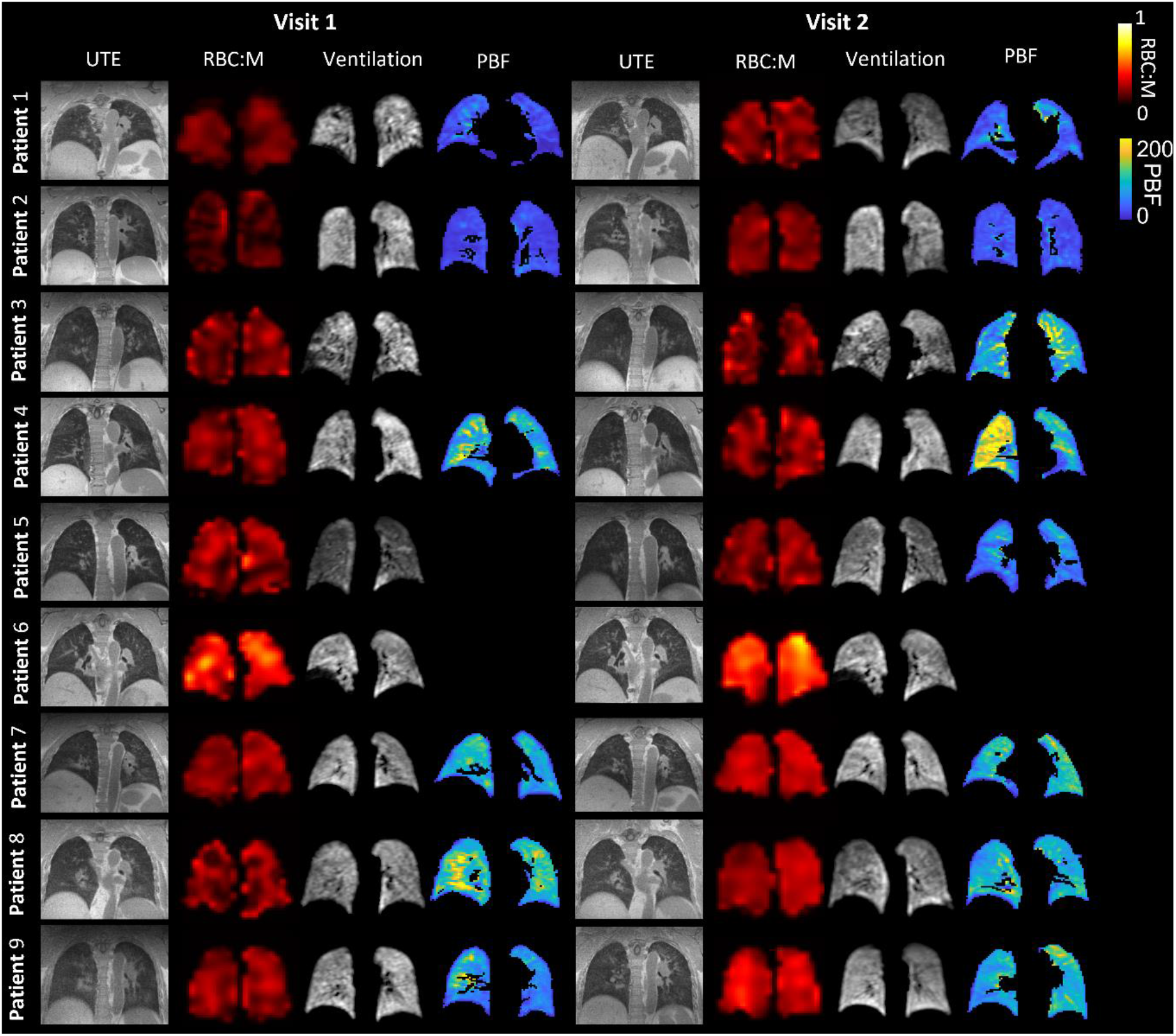
Example ultra-short echo time (UTE) images, RBC:M maps, ^129^Xe ventilation images and maps of pulmonary blood flow at visit 1 and visit 2, for each patient. The white arrow shows a segmental perfusion defect visible at visit 1 which improves at visit 2. UTE: ultra-short echo time. RBC: red blood cell. M: membrane. PBF: pulmonary blood flow.

**Figure 4:**
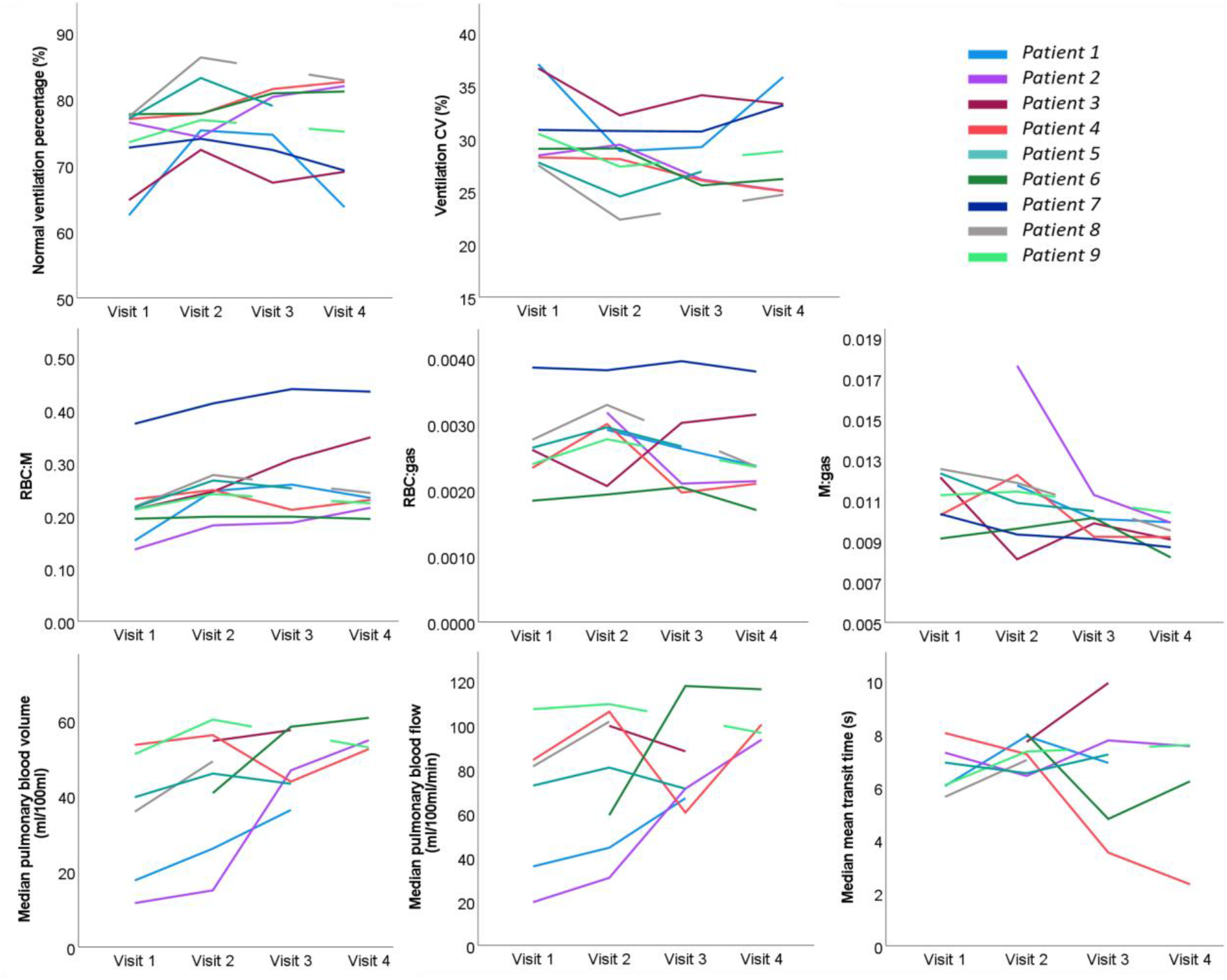
Spaghetti plots of ventilation, dissolved phase xenon and DCE lung perfusion metrics at visits 1-4. VDP: ventilation defect percentage. CV: coefficient of variation of lung ventilation. RBC: red blood cell. M: membrane.

### ^129^Xe MRI

#### Ventilation

At visit 1, small ventilation defects were visible in the lung periphery in four patients (1, 3, 4 and 6). No other patients had visible lung ventilation defects. At visit 2 and 3, the ventilation defects observed in patients 1, 3, 4 and 6 had improved, with small defects still visible particularly in patient 3. At visit 4, small peripheral ventilation defects were observed in patients 1, 3 and 6 see Figures 4 and Supplementary Figure 1. At visit 1, VDP was 1.6 (0.6-3.9)%, at visit 2 VDP was 1.3(0.7-2.6)%, at visit 3 VDP was 1.2(0.4-2.1)% and at visit 4 VDP was 0.8(0.4-3.7)%.

Quantitative metrics of ventilation improved at visits 2, 3 and 4 compared to visit 1, however this was not statistically significant after adjustment for multiple corrections, see Table 2.

#### DW-MRI (alveolar microstructure)

Median ADC and Lm_D_ at each visit are reported in Table 2. No significant longitudinal changes in ADC and Lm_D_ were seen between visits. Median ADC and Lm_D_ were within the interquartile range (IQR) of age and sex matched control data (age and sex matched control data: median ADC=0.0360 cm^2^s^-1^, IQR=0.005cm^2^s^-1^; Median Lm_D_ =289µm, IQR=27µm) at all visits.

#### Dissolved xenon (gas exchange)

The global RBC:M ratio significantly increased at visit 2 when compared to visit 1 (p_adj_=0.023). RBC:M at visit 1 was 0.22(0.15-0.37) and at visit 2 was 0.25(0.18-0.41). No subjects showed a decrease in RBC:M at visit 2 when compared to visit 1, see Figures 4 and 5. RBC:M at visits 3 and 4 were of 0.25(0.19-0.44) and 0.23(0.19-0.44) respectively. At visits 3 and 4, some patients showed continued improvement (see Figures 3 and 4), while others maintained an abnormal RBC:M during the 25-51 week period. There were no significant changes between visits 2, 3 and 4.

**Figure 5:**
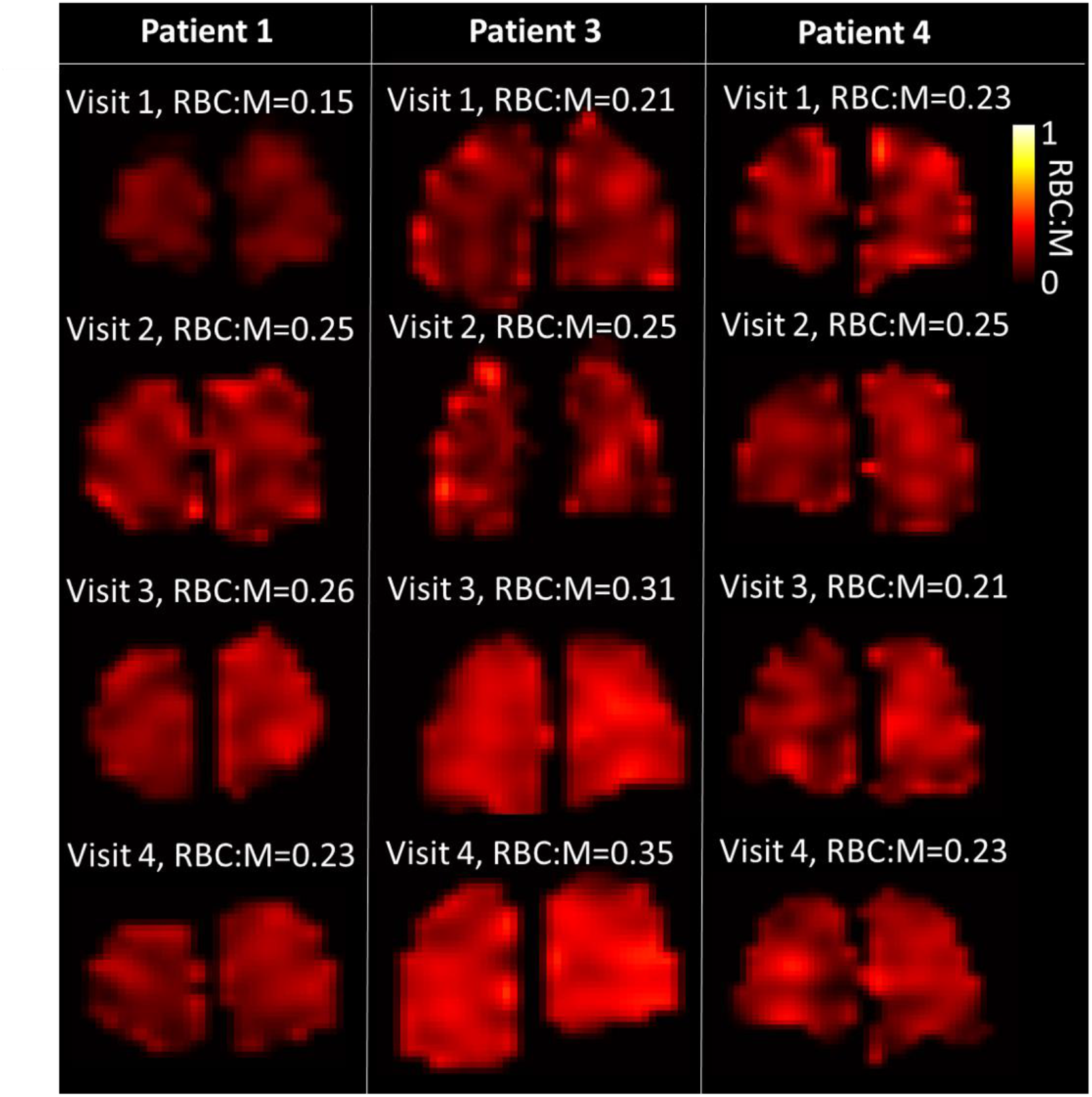
Lung RBC:M maps in three patients with four MRI visits at 6, 12, 25 and 50 weeks after hospital admission. Mean RBC:M at each visit is shown. RBC: red blood cell. M: Membrane.

Figure 6 shows box-plots of the RBC:M, RBC:gas and M:gas for patients at each visit, with reference box plots of age and sex matched control data (control RBC:M median=0.39, IQR=0.13; RBC:gas median=0.0034, IQR=0.0006; M:gas median=0.0088, IQR=0.0021). The number of patients who had RBC:M below the IQR of the age and sex matched healthy volunteers was 8/9 at visit 1, 7/9 at visit 2, 5/7 at visit 3 and 6/8 at visit 4.

**Figure 6:**
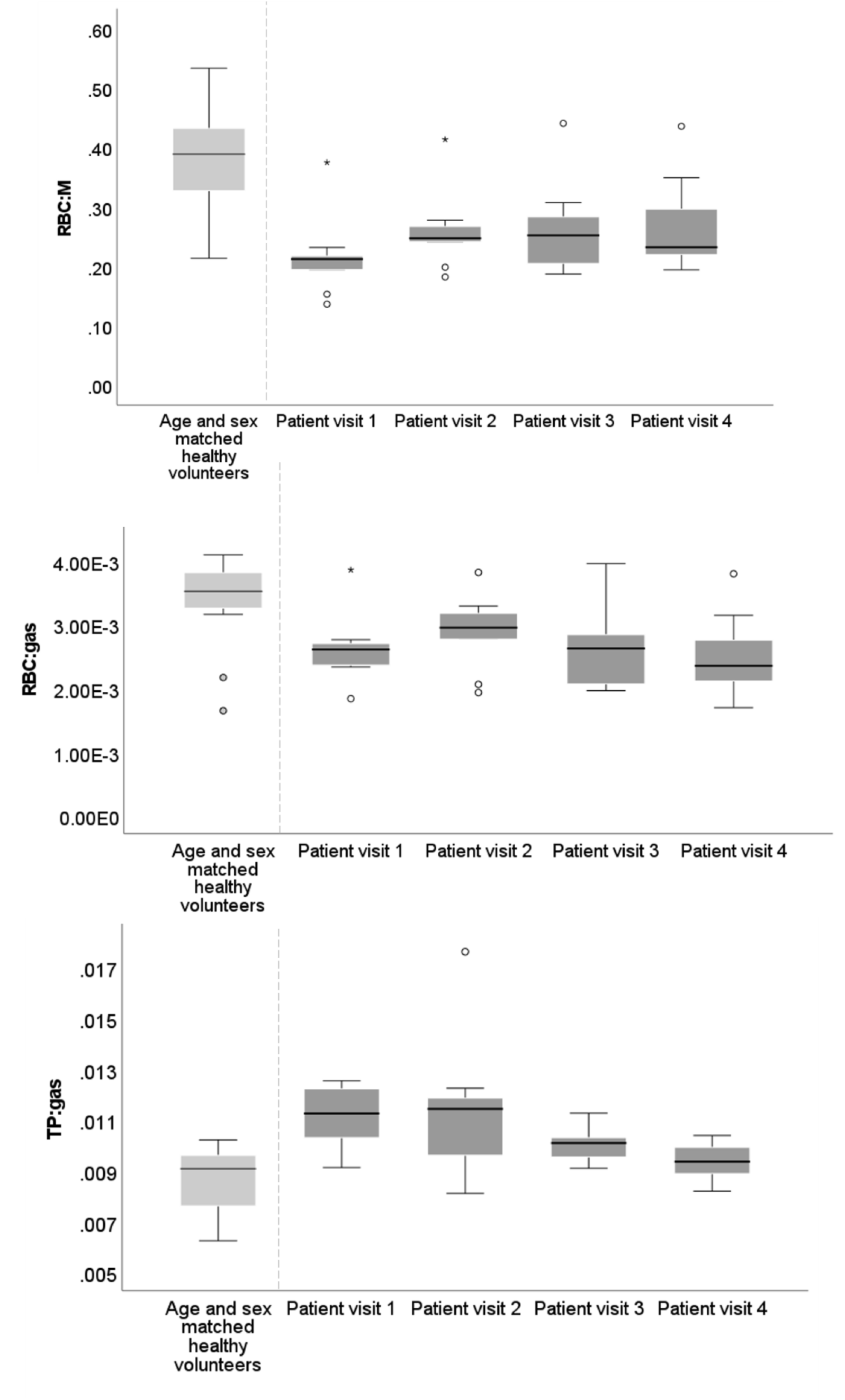
Box plots of gas transfer ratios from patients at visit 1, visit 2 and visit 3 as well as metrics from an age and sex matched healthy cohort. RBC: red blood cell. TP: tissue/plasma. o denotes data >1.5 IQR, ★ denotes data >3 IQR.

M T_2_^*^ also showed a significant longitudinal decrease across visits, with lower M T_2_^*^ at visit 4 compared to visits 1, 2 and 3 (p_adj_=0.023, p_adj_=0.023, p_adj_=0.047) and between visits 1 and 3 (p_adj_=0.031), see Table 2. No other significant changes in RBC or M T_2_^*^were seen, see the online supplement Supplementary Figure 2.

### ^1^H MRI

#### Structural changes

The UTE image of patient 3 showed abnormal linear parenchymal changes at visit 1, which improved but remained abnormal at visits 2 and 3 and were resolved at visit 4. Patients 2, 6, 7 and 8 showed air trapping on their UTE image at visit 1, which resolved at visit 2 for patients 6, 7 and 8. Patient 2 continued to have air trapping present at visits 3 and 4. The UTE images of patients 1, 4, 5 and 9 were normal at all visits, see the online supplement, Supplementary Table 2.

#### DCE (Perfusion)

Patient 1 showed a segmental perfusion defect at visit 1 which was resolved at visit 2. No other patients showed any substantial regional perfusion defects. Median pulmonary blood volume and flow increased in all patients (n=6) at visit 2 compared to visit 1, however the increase was not statistically significant. For the 6 patients with DCE MRI at visit 1 and 2, pulmonary blood volume was 37.8 (11.7-53.5) ml/100ml at visit 1 and 47.6 (15.0-60.2)ml/100ml at visit 2, and pulmonary blood flow was 76.9 (19.6-107.2)ml/100ml/min at visit 1 and 91.1 (30.7-109.5)ml/100ml/min at visit 2, see Figure 4.

#### Correlation of RBC:M and DCE metrics

The analysis of covariance showed a statistically significant correlation between RBC:M and pulmonary blood volume within patients, across visits 1-4. Correlation coefficient=0.49, p=0.029. There was also a statistically significant within patient correlation between RBC:M and mean transit time, correlation coefficient=0.455, p=0.032, see online supplement Supplementary Figure 3. No statistically significant correlations between RBC:M and pulmonary blood flow found.

#### PFTs

data was available for 6/9 patients at visit 1, 6/9 patients at visit 2, 7/7 patients at visit 3 and 7/8 patients at visit 4, all Z-scores and % predicted data are shown in Table 3. There was a median of 0 days (mean: 2.8 days, range: 0-23 days) between MRI and PFT tests.

**Table 3.**
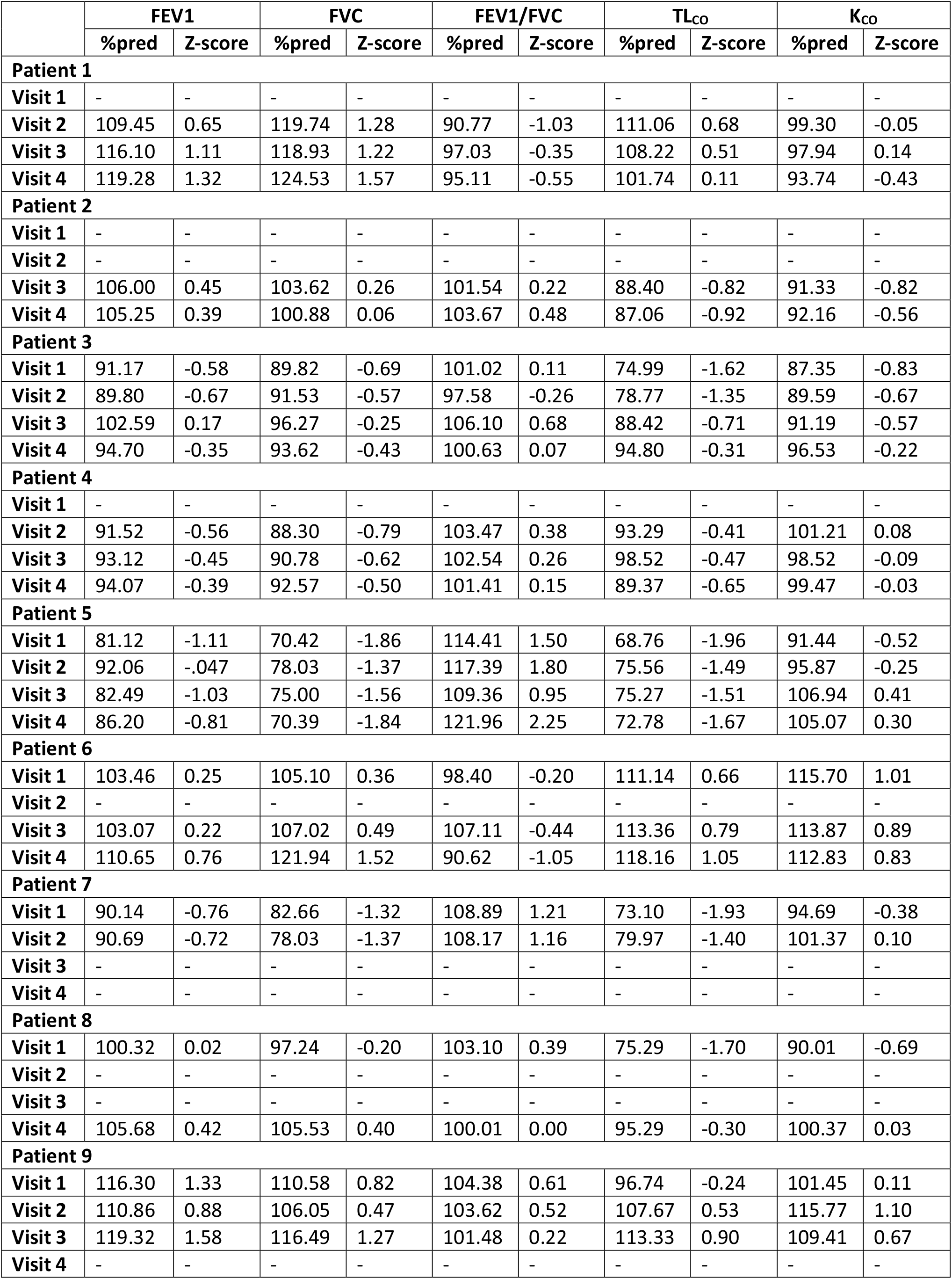
All available PFT data (% predicted and Z-score) at visits 1, 2 and 3.

TL_CO_ Z-score (TL_CO_-Z) was -1.66 (−1.96, 0.66) at visit 1, -0.88 (−1.49, 0.68) at visit 2, -0.47 (−1.51, 0.90) at visit 3 and -0.31 (−1.67,1.05) at visit 4. 3/6 patients had abnormal TL_CO_-Z(<1.64) at visit 1. No patients had abnormal TL_CO_-Z at visit 2 or 3. 1 patient had abnormal TL_CO_-Z at visit 4, see Supplementary Figure 4.

One patient (patient 5) had abnormally low FVC at visit 1 and visit 4. No other PFT metrics were outside abnormal at any visits.

The analysis of covariance showed a statistically significant correlation between RBC:M and TL_CO_ Z-score within patients across visits 1-4, correlation coefficient=0.64, p<0.001.

## Discussion

This study has used a comprehensive multinuclear MRI protocol to assess pathophysiological pulmonary changes in patients who were hospitalised with COVID-19 up to a year after hospitalisation. The preliminary findings demonstrate that these patients had impaired gas transfer (RBC:M), however measures of lung microstructure (ADC and Lm_D_) were normal. Four of nine patients had small ventilation defects at 6 weeks which largely resolved by 25 weeks. Despite significant increases in ^129^Xe gas transfer (RBC:M) at 12 weeks when compared to 6 weeks, median^129^Xe gas transfer in these patients remains lower than expected at 12-51 weeks, compared to an age and sex matched healthy cohort. These initial results indicate that some COVID-19 patients show continued abnormalities in ^129^Xe gas transfer at 12-51 weeks after hospitalisation, while others demonstrate consistent improvement across the same time frame, with 2 patients within the IQR of normal age and sex matched patients at 51 weeks.

Li *et al*. reported a significantly lower mean RBC:M values of 0.279±0.04 at discharge in patients hospitalised with COVID-19 pneumonia when compared to healthy volunteers[11] while Grist *et al*. reported a significantly lower mean RBC:M of 0.3±0.1 at 24 weeks after discharge[12] and a mean RBC:M of 0.31±0.10 in post-hospitalised patients 21±10 weeks after infection[13]. Although not directly comparable due to differences in imaging parameters, our findings are in accordance with the reporting of significantly lower RBC:M values between hospital discharge and 24 weeks post discharge. In addition, the inclusion of data from age and sex matched healthy controls demonstrates that these changes are not due to age or sex differences between controls and patients in this study. RBC:gas and M:gas did not show significant longitudinal change once adjusted for multiple comparisons, implying that the change in RBC:M was a combined effect of changes in both M and RBC. A significant reduction in M T_2_^*^ at visit 2 was also found. The physiological mechanisms behind changes in M T_2_^*^ are not well established and for further discussion, see the online supplement.

In this work we demonstrate that this patient cohort has impaired xenon gas transfer (RBC:M), with reference data from a healthy age and sex matched cohort presented, and that changes in xenon gas transfer correlate positively and significantly with clinical markers of lung function (TL_CO_Z) and lung perfusion metrics (pulmonary blood volume and mean transit time). All patients with DCE data available showed an increase in regional pulmonary blood flow and volume between visit 1 and 2, despite only one having a substantial perfusion defect. This may indicate microvascular improvements at 12 weeks, and that microvascular recovery may be partially driving changes in RBC:M in these patients. In parallel a concomitant reduction in M signal due to resolution of post-infection endothelial inflammation could contribute to the increase in RBC:M with time.

In this study, most patients (7/9) did not report significant breathlessness at visit 2 (12 weeks), despite lower RBC:M than the control reference data. The two patients that reported breathlessness at visit 2 had the two lowest RBC:M values at that visit. It is possible that decreased RBC:M is measuring sub-clinical changes in gas transfer. This may inform the interpretation of other studies showing decreased RBC:M in a breathless population, however larger studies in symptomatic patients are needed to further investigate links between RBC:M and breathlessness or other post-COVID symptoms.

Median patient ADC and Lm_D_ were within the age and sex matched reference range at visits 1 and 2[23], with no significant change at visit 2, indicating normal lung microstructure in 9 patients who had COVID-19 but did not have signs of interstitial lung damage on structural imaging. This paper specifically excluded patients with signs of interstitial lung damage, however it has been reported that at 6 weeks after hospitalisation 4.8% of patients had radiological changes compatible with early interstitial lung damage[36], and DW-MRI has been shown previously to be sensitive to changes in alveolar geometry in patients with interstitial lung damage[24]. Therefore, further work using DW-MRI in patients with pulmonary fibrosis on CT due to COVID-19 is warranted to evaluate changes in the lung microstructure of these patients and is the subject of an ongoing study (UKILD)[20].

Minor ventilation heterogeneity and defects were present in this cohort shortly after acute illness, which improved over time, consistent with the previous findings by both Grist *et al*.[12] and Li *et al*.[11]. *Li et al*., reported a ventilation defect percentage (VDP) of 5.9% (range: 1.6-9.9%) in patients discharged after hospitalisation due to COVID-19 (14-34 days after discharge) which was significantly higher than in healthy volunteers (3.7%). *Grist et al*., reported well ventilated lungs in patients after having had COVID-19 (mean time after discharge: 24 weeks) although the imaging used to assess ventilation in that study was of poorer spatial resolution so may have missed mild ventilation heterogeneity. Overall, our study and the findings from previous literature suggest it is unlikely that impaired lung ventilation is the primary cause of ongoing symptoms after the acute stage of COVID-19 and that the pathophysiology is not primarily of the airways.

The main limitation of this study is the limited number of participants which was largely due to the challenging nature of recruiting patients for scanning directly after a recent hospitalisation due to COVID-19, and in addition not all patients had DCE lung perfusion imaging or PFTs at all examinations. The numbers recruited limit correlations with symptoms, activity and lung function and also the statistical tests used to test for change. A potential source of bias in this study is that 5 patients who were potentially eligible for the study were excluded due to chest size exceeding the size of the xenon MRI coil.

This paper has focused on the longitudinal analysis of the global whole lung metrics derived from the functional imaging techniques. Work in progress entails regional analysis of the individual functional techniques and investigation of the anatomical distribution and their regional correlation.

In conclusion, the initial results from this ongoing longitudinal study show that 6 weeks after hospital admission with COVID-19 pneumonia, patients demonstrated impaired ^129^Xe gas transfer (RBC:M), but normal lung microstructure (ADC, Lm_D_). Small ventilation defects and heterogeneity present in some patients largely resolved in the 6-25 week period. Improvements in an individual’s RBC:M correlated with an increase in an individual’s lung perfusion and TL_CO_z. At 12 week follow up, improvements in xenon gas transfer (RBC:M) were seen, though RBC:M remained abnormal for the majority of patients. At 25-51 week follow up, patient RBC:M changes were heterogenous, with some patients showing continued improvement whilst for others, RBC:M remained abnormal.

We believe this to be the first follow up study of such patients with such an extensive range of functional lung imaging techniques and the findings demonstrate the sensitivity and complementary nature of functional MRI to follow up post COVID lung pathophysiology in a clinical setting.

## Data Availability

All data produced in the present study are available upon reasonable request to the authors

## Acknowledgements

JMW Medical Research Council grant “Expansion of state-of-the-art MR imaging infrastructure for pulmonary disease stratification: POLARIS” MR/M008894/1

GSK and GE for investigator led grant funding

AART was supported by a BHF Intermediate Clinical Fellowship (FS/18/13/33281)

## Supplementary material

## Methods

### Dissolved phase xenon acquisition and analysis

The spectroscopic imaging method for the first visit of subjects 1 and 2 followed the 4-echo flyback 3D radial technique[24]. For the remaining visits and all other subjects, an updated version of the imaging method was implemented. The main improvements being: (i) the implementation of a frequency-tailored RF excitation pulse that excites the gas phase with 1% of the flip angle on tissue and blood, to allow a more efficient sampling time by removing the interleaved RF excitation between gas and dissolved phase and increase the number of radial projections from 332 to 934, which therefore decreased the radial undersampling; (ii) the TR was shortened to 15 ms and the flip angle reduced from 40 to 22 degrees to match the imaging parameters recommended by the ^129^Xe MRI clinical trials consortium[13]; (iii) the calibration spectrum required to estimate the frequency positions and transverse relaxation times (T_2_^*^) of each resonance was inserted at the start of the imaging sequence. These estimates were used as prior information to improve the chemical shift reconstruction and were derived by performing a triple Lorentzian fit of the data in the frequency domain[14]. This latter addition removed the use of a separate calibration dose and sequence in the imaging protocol. The remaining imaging parameters (FOV, matrix size, BW, echo times) stayed the same and are shown in the supplementary table 1.

A correction factor for the visit 1 of subjects 1 and 2 RBC:M ratio was applied to account for the different TR and flip angle between the two implementations of the dissolved phase imaging technique. The correction factor was estimated using images acquired using both techniques on 6 healthy volunteers. RBC:Gas and M:Gas ratio were discarded for these patients (n=2), as a reliable correction factor was not found.

### Linear binning methodology for lung ventilation imaging

A linear binning approach[38] based on [39] was used to compute a 4 level grading of the ventilation images as follows: (1) pre-computed estimations of the lung cavity and major airways[40] were manually edited; (2) the N4 bias field correction filter[19] was applied to the ventilation images, where the ventilated-region mask was supplied by an in-house-developed deep-learning tool; (3) finally, normalisation by the mean signal inside the lung cavity and linear binning were performed with the lower bins being the defect and low ventilation regions. Coefficient of variation (CV) of ventilation images was calculated from the 3D ventilated volume as a measure of ventilation heterogeneity[41].

### UTE imaging

A correction for receiver coil signal non-uniformity (PURE) was performed retrospectively on all images using GE Orchestra software.

## Discussion

A significant reduction in M T_2_^*^ at visit 2 was found. The physiological mechanisms behind changes in M T_2_^*^ are not well established. An increased M T_2_^*^ at visit 1 may be due to residual inflammation increasing the thickness of interstitial tissue, potentially resulting in a decrease in microscopic field inhomogeneity effects in the interstitium. However, further work is warranted to determine this relationship.

**Supplementary table 1.** typical imaging parameters used for MRI acquisition

|  | <sup>129</sup>Xe ventilation | <sup>129</sup>Xe DW-MRI | Dissolved phase <sup>129</sup>Xe | <sup>1</sup>H UTE | <sup>1</sup>H VFA | <sup>1</sup>H DCE |
| --- | --- | --- | --- | --- | --- | --- |
| Sequence | 3D bSSFP sequence | 3D SPGR multiple b-value with elliptical-centric phase encoding | 4-echo flyback 3D radial technique | 3D SPGR radial sequence | 3D SPGR | 3D SPGR |
| Acquisition matrix | 100x100 | 64 × 52 | 32x32x32 | 256 x 256 x 256 | 200x80 | 200x80 |
| Field of view (cm<sup>2</sup>) | 48 | 48 | 40 | 35 | 48 | 48 |
| Slice thickness (mm) | 10 | 15 | 0.55 | 1.37 | 4 | 10 |
| TE (ms) | 2.17 | 14.0 |  | 0.08 | 0.9 | 0.69 |
| TR (ms) | 6.52 | 17.4 | 15 | 2.9 | 2.85 | 2.08 |
| FA (°) | 10 | 3 | 22 | 4 | 2°, 4°, 10°, 30° | 30° |
| Temporal acquisitions | - | - | - | - | - | 48 |
| Temporal resolution (s) | - | - | - | - | - | ~0.5 |
| SENSE factor | - | - | - | - | - | 2 |
| Xenon dose | 500ml | 550ml | 1000ml | - | - | - |
| Bandwidth (Hz/pixel) | 62.97 | 54.22 | 31.25 | 1953.10 | 488.28 | 976.56 |
| Respiratory state | FRC + 1L bag of gas (<sup>129</sup>Xe and N<sub>2</sub> mixture) | FRC + 1L bag of gas (<sup>129</sup>Xe and N<sub>2</sub> mixture) | FRC + 1L bag of gas (<sup>129</sup>Xe) | Expiration, prospectively gated free breathing | Expiration breath hold | Expiration breath hold followed by shallow breathing |
| Additional parameters |  | b = 0, 12, 20, 30 s/cm<sup>2</sup><br>diffusion time (Δ) = 8.5 ms | ΔTE = 0.7 |  |  |  |

**Supplementary table 2.** UTE radiological assessment.

| UTE radiological assessment |  |  |  |  |
| --- | --- | --- | --- | --- |
|  | Visit 1 | Visit 2 | Visit 3 | Visit 4 |
| **Patient 1** | Normal | Normal | Normal | Normal |
| **Patient 2** | Air trapping | Air trapping | Air trapping | Very minor air trapping |
| **Patient 3** | Abnormal linear parenchymal changes and air trapping. | Parenchymal changes are present but improved. | Parenchymal changes are present but further improved. | Normal |
| **Patient 4** | Normal | Normal | Normal | Normal |
| **Patient 5** | Normal | Normal | Normal | Normal |
| **Patient 6** | Air trapping | Normal | Normal | Normal |
| **Patient 7** | Air trapping | Normal | - | Normal |
| **Patient 8** | Air trapping | Normal | - | Normal |
| **Patient 9** | Normal | Normal | Normal | - |

